# Enhanced 40 and 80 Hz Auditory Steady State Responses in Idiopathic Tinnitus

**DOI:** 10.1101/2024.03.02.24303654

**Authors:** Zahra Ghasemahmad, Saeid Mahmoudian, Daniel Gavazzi, Shohreh Jalaei, Saeid Farahani

**Author notes:** Corresponding author: Zahra Ghasemahmad, Ph.D.

## Abstract

**Objectives:** This study aimed to investigate changes in auditory processing using auditory steady state responses (ASSR) in patients with idiopathic tinnitus.

**Method:** 19 tinnitus patients and 23 control subjects without tinnitus were examined with multiple ASSR. Three modulation frequencies of 20, 40, and 80 HZ were tested and the steady state responses were compared between tinnitus and control group. Further, the thresholds in ipsi- and contralateral side to ear with tinnitus were compared.

**Results:** Our findings showed no significant difference in ASSR thresholds in ipsi- and contralateral side in tinnitus patients. However, we observed enhanced ASSRs at 40 and 80 Hz modulations in patients with idiopathic tinnitus compared to no-tinnitus control subjects.

**Conclusions:** The results of this study suggest possible sensory deficits along higher order auditory regions in patients with idiopathic tinnitus. Further, our data indicates a bilateral involvement of auditory pathway in these regions in patients with lateralized tinnitus.

## INTRODUCTION

Tinnitus is the disturbing sensation of sound in one ear or both without any external stimulation and can cover 4.4-15% of any population at any age [1]. 5-15% of society experience chronic sensation of tinnitus [2]. Although tinnitus has been reported and studied as a symptom for years, neuroscience theories about tinnitus are very young and most of the findings are based on the results from failed attempts. Though hearing loss has been recognized as the primary and main reason for tinnitus, other cases with normal hearing and no diagnosed pathological reason have also been reported [3]. In fact, in over 50% of patients referred to tinnitus clinics, the physiological and electrophysiological examinations have failed to diagnose the underlying cause of tinnitus and thus, have been classified as idiopathic [4]. Since tinnitus is merely a symptom and its control and management rely on diagnosing its underlying cause, the necessity of localizing the involved regions in patients with idiopathic tinnitus is undeniable.

Tinnitus studies have identified various brain regions with abnormal activities along the auditory pathways. For instance, using positron emission tomography (PET) [6] and functional magnetic resonance imaging (fMRI) [7–10] have shown changes in auditory cortex in tinnitus patients while other studies have emphasized the role of inferior colliculus (IC) [11, 12] and medial geniculate body (MGB) [13, 14]. However, comparing and generalizing the findings between these studies is difficult [5] for a few reasons. First, as tinnitus is a symptom rather than a disease, including patients with various etiologies might result in different localization of tinnitus regions as well as diverse treatment plans. Second, since in most tinnitus studies subjects suffer from hearing loss, it is not clear whether the abnormal neural activity along the auditory pathway is the result of tinnitus or hearing loss. Lastly, employing different techniques for examining different brain regions makes drawing comparisons between studies harder. Thus, such reasons call for the need to employ one technique and apply harder inclusion criteria to subjects to eliminate possible factors that could influence the localization of tinnitus sites or affected regions.

A possible technique for localizing the source of tinnitus is the auditory steady state response (ASSR). ASSRs are event-related potentials (ERPs) that provide means to evaluate the integrity of auditory pathways with minimal demands and interventions [15, 16]. ASSR technique allows simultaneous measurement of the responses originated in various parts of the auditory pathway in both ears and all carrier frequencies (CF) [17, 18].

ASSRs rely on continuous modulated pure tones with different carrier frequencies, usually octave frequencies of 500–4000 Hz, for measuring activity along auditory pathway [19]. Depending on modulation frequency (MF), various brain regions have been identified to be responsible for generating these responses. Studies have suggested the involvement of regions such as brainstem, midbrain, and auditory cortex responses to 80, 40, and 20 Hz MF, respectively [20,21]. Even though the ASSR thresholds might not be reliable sources for estimating hearing thresholds in normal hearing adults [22–24], its ability to test multiple frequencies simultaneously, objectivity, noninvasiveness, and evaluate various sites along auditory pathway [25] can be an invaluable tool in diagnosis of auditory deficits such as tinnitus.

The goal of the current study was to examine ASSR thresholds in various modulation frequencies in adults with idiopathic tinnitus and no-tinnitus control. Using an age and gender matched group allowed us to use a controlled approach to evaluate such cortical and subcortical thresholds in tinnitus subjects. We evaluated these responses in the two groups for all modulation frequencies and then compared these results with those in no-tinnitus control. We further assessed the response magnitudes in ipsi- and contralateral side relative to the ear with tinnitus sensation in patients with idiopathic tinnitus to understand the impact of the tinnitus laterality on the possible changes in these responses.

## METHOD

### Ethical considerations

The study was approved by the local ethics committee at Iran University of Medical Sciences and Tehran University of Medical Sciences and informed consent was obtained from each participant.

### Subjects

24 normal hearing adults (15 females and 9 males, 25-40 years old) and 19 tinnitus patients (7 females and 12 males, 25-45 yrs old) with hearing thresholds at or better than 20 dB HL at 6 octave frequencies: 250, 500, 1000, 2000, 4000 and 8000 Hz participated in this study. The ASSRs from the control group were used from a previous publication [22]. All tinnitus patients were selected from the population of tinnitus subjects referred to the tinnitus research center at the otolaryngology department of the Hazrat Rasool Akram (PBUH) Educational, Research, and Treatment Hospital in Tehran, Iran. Only subjects without an underlying condition were included in the study. The inclusion and exclusion criteria can be viewed in Table 1.

**Table 1:**
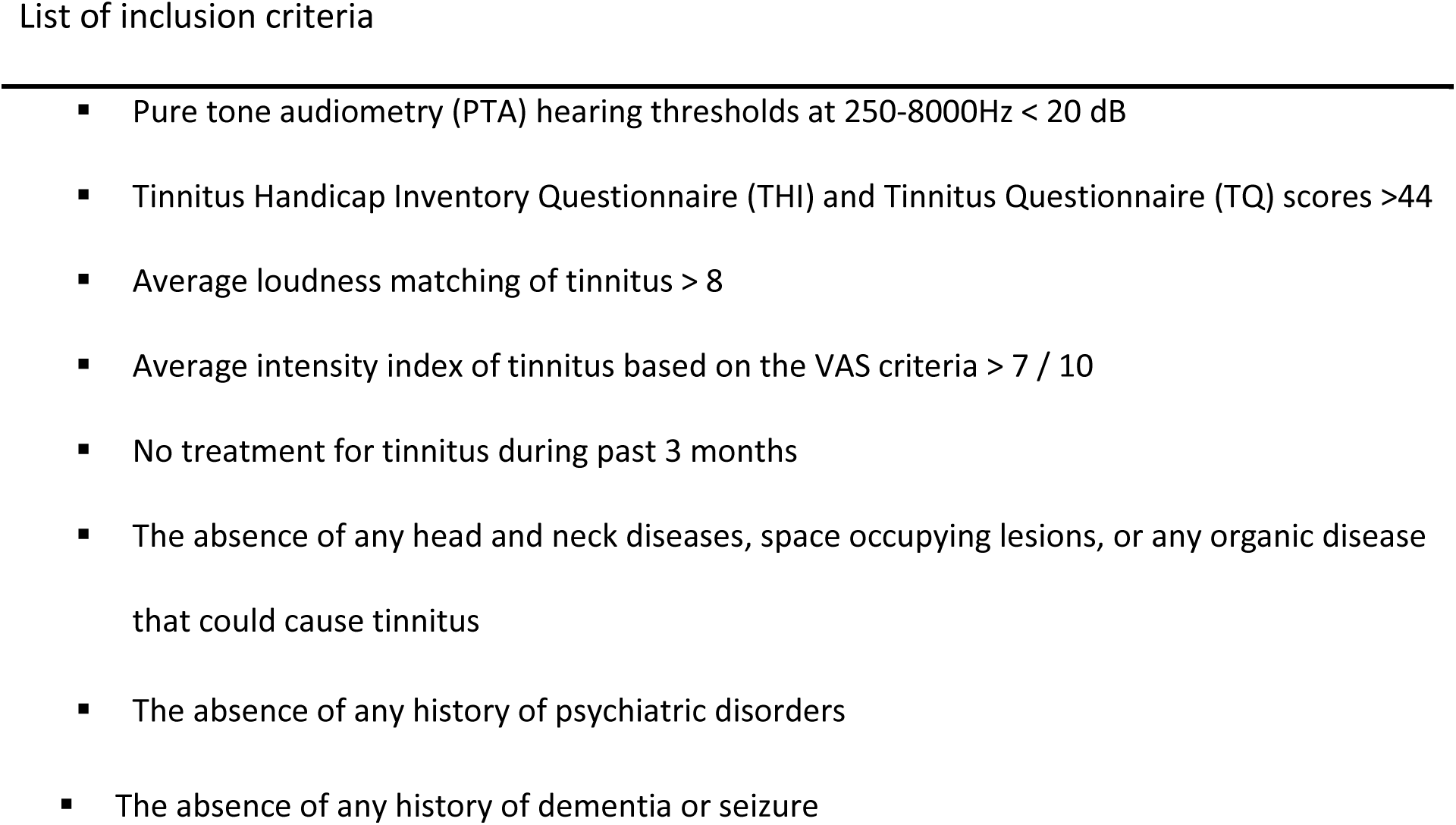
Summary of the criteria for inclusion or exclusion from the study.

### Experimental protocol

#### ASSR

ASSR was performed in a soundproof room, and thresholds were measured based on the Hughson-Westlake approach (10 down-5 up). Air-conducted stimuli were introduced into each ear through an ER-3A insert earphone (Etymotic Research). Stimuli were generated and presented by the CHARTR EP system (version 5.3, GN Otometrics) and presented to both ears simultaneously. All the stimuli were 100% amplitude-modulated (AM) and 20% frequency–modulated (FM). Response confidence, a measure of signal to noise ratio and the chance of finding a response, was kept at 95%. The response confidence in this system is calculated based on magnitude-squared coherence (MSC) algorithm to detect the probability of response presence relative to surrounding noise [26]. ASSRs were performed at four carrier frequencies of 500, 1000, 2000, and 4000 Hz and intensity range of 10-80 dB SPL. The detailed modulation rate choice for each combination of MF and CF are as previously reported here [22].

During each session, subjects relaxed in a supine position on a comfortable bed. Participants were asked to sleep or relax during the 80 Hz multiple ASSR testing. However, as 20 Hz and 40 Hz ASSR responses could be affected by sleep participants were engaged in performing simple math problems to prevent them from falling sleep.

Responses were recorded via three scalp electrodes: the active and ground electrodes on the upper and lower forehead respectively, and the reference electrode on the nape. The impedance of the electrodes was periodically checked during recordings to maintain low impedance and a good connection (less than 3 KΩ). To minimize movement related artifacts in head and body that could affect the signal to noise ratio, the participants were asked to stay relaxed and motionless during the experiment.

### Statistical analysis

Due to the smaller sample size, nonparametric tests were used to analyze ASSRs. For comparing the ASSR thresholds between ipsi- and contralateral sides in tinnitus patients at each combination of carrier and modulation frequencies, a Wilcoxon signed rank test was used. Results were considered significant if the *p-value* was less than 0.05. The obtained ASSR thresholds for tinnitus patients and no-tinnitus control were analyzed and compared using Kruskal Wallis test and were compensated for multiple comparisons using a Bonferroni correction. Thus, the *p-values* in this comparison were divided by the number of comparisons made, making *p-values* significant at 0.025. All Statistical analysis was performed using IBM SPSS Statistics version 27. All graphs were generated using custom written python code.

## RESULTS

PTA and ASSR thresholds were obtained in 19 tinnitus patients and 24 no-tinnitus control. Tinnitus was lateralized in all patients (9 subjects in the right and 10 in the left ear). All tinnitus patients were selected based on the inclusion criteria summarized in Table 1.

We first sought to compare the ASSR thresholds in the ipsi- and contralateral side relative to the ear with tinnitus sensation in all tinnitus subjects. The modulation frequencies of 20 and 40 Hz ASSRs are believed to originate mainly in midbrain and cortex [27], and the auditory pathway in these regions mostly processes the contralateral input [28]. As a result, the right ear responses were registered for the left ear and the left ear’s responses for the right ear for these two modulation frequencies. The ipsi- and contralateral ASSRs were recorded respectively.

The mean and standard deviation of these responses for each combination of MF and CF are summarized in Table 2. As shown in this table, the highest (worst) ASSR thresholds are observed for both ipsi- and contralateral sides in 20 Hz MF, with the highest mean ± standard deviation recorded at 4000 Hz CF (54.2 ± 14.3 dB SPL in ipsilateral ear vs 57.9 ± 11.9 dB SPL in contralateral side). The responses in other MFs showed lower average values consistent with previously reported thresholds in normal hearing adults [22]. The ASSR responses in both ipsi- and contralateral side showed lowest thresholds at 2000 HZ compared to other carrier frequencies in most MFs. Among all MFs, 40 Hz showed the lowest thresholds for most carrier frequencies (See Table 2).

**Table 2:**
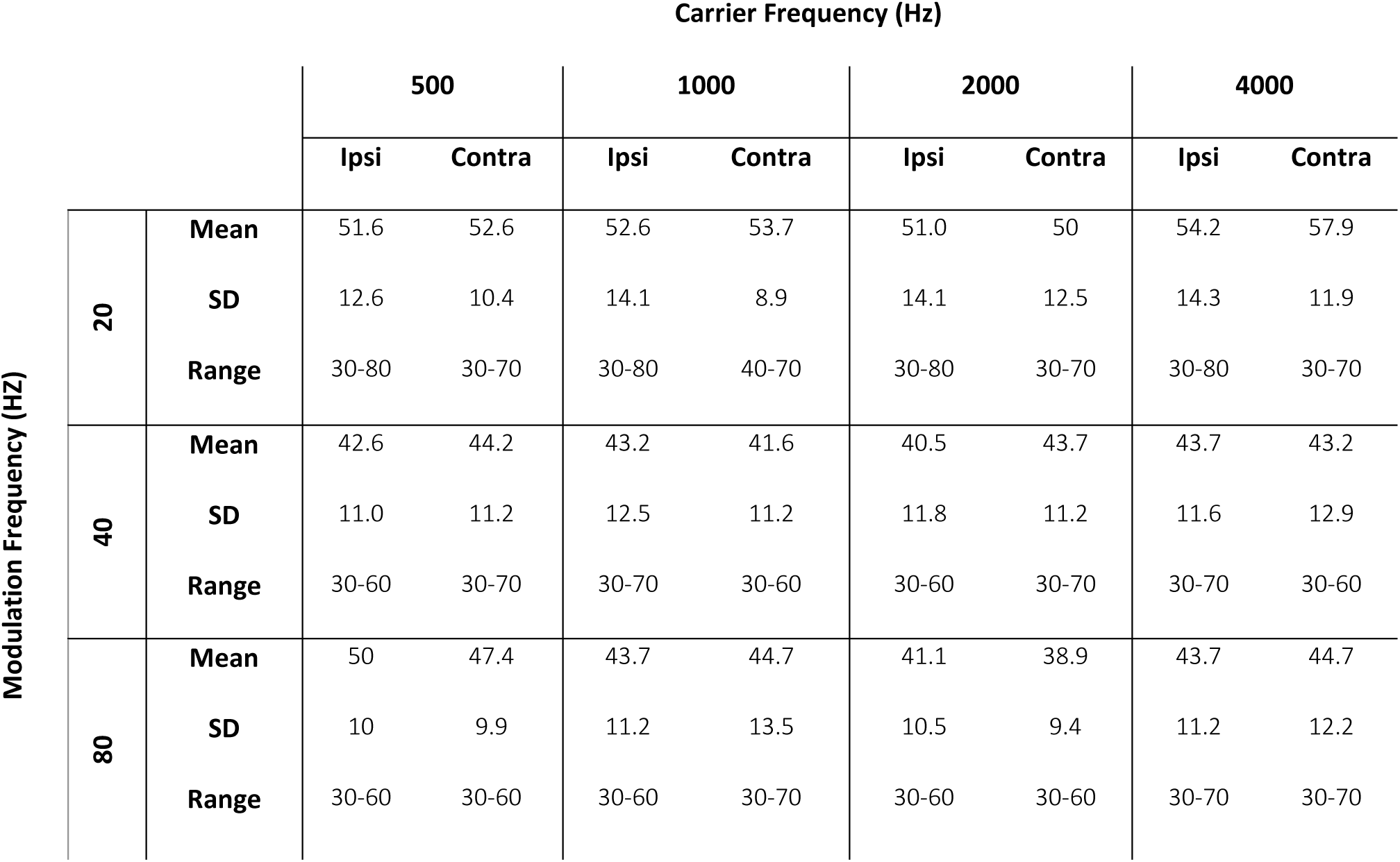
Mean values (Mean), standard deviation (SD) and range (Range) of Auditory Steady State Responses (ASSR) in dB SPL for modulation frequencies of 20, 40, 80Hz and in carrier frequencies 500-4000Hz in ipsi- and contralateral ear to tinnitus ear in patients with subjective idiopathic tinnitus, n=19 subjects.

The single data points of ipsi- and contralateral ASSR thresholds for each combination of these thresholds are shown in Fig. 1. As evident from these scatterplots, the ipsi- and contralateral ASSRs showed a linear relationship between the two sides. Further, comparing the ASSR thresholds in ipsi- and contralateral side in tinnitus patients in each combination of MF and CF showed no significant differences in ASSRs obtained in the two sides relative to tinnitus perception side (Wilcoxon signed rank test; see statistical analysis results in Table 3).

**Fig. 1.**
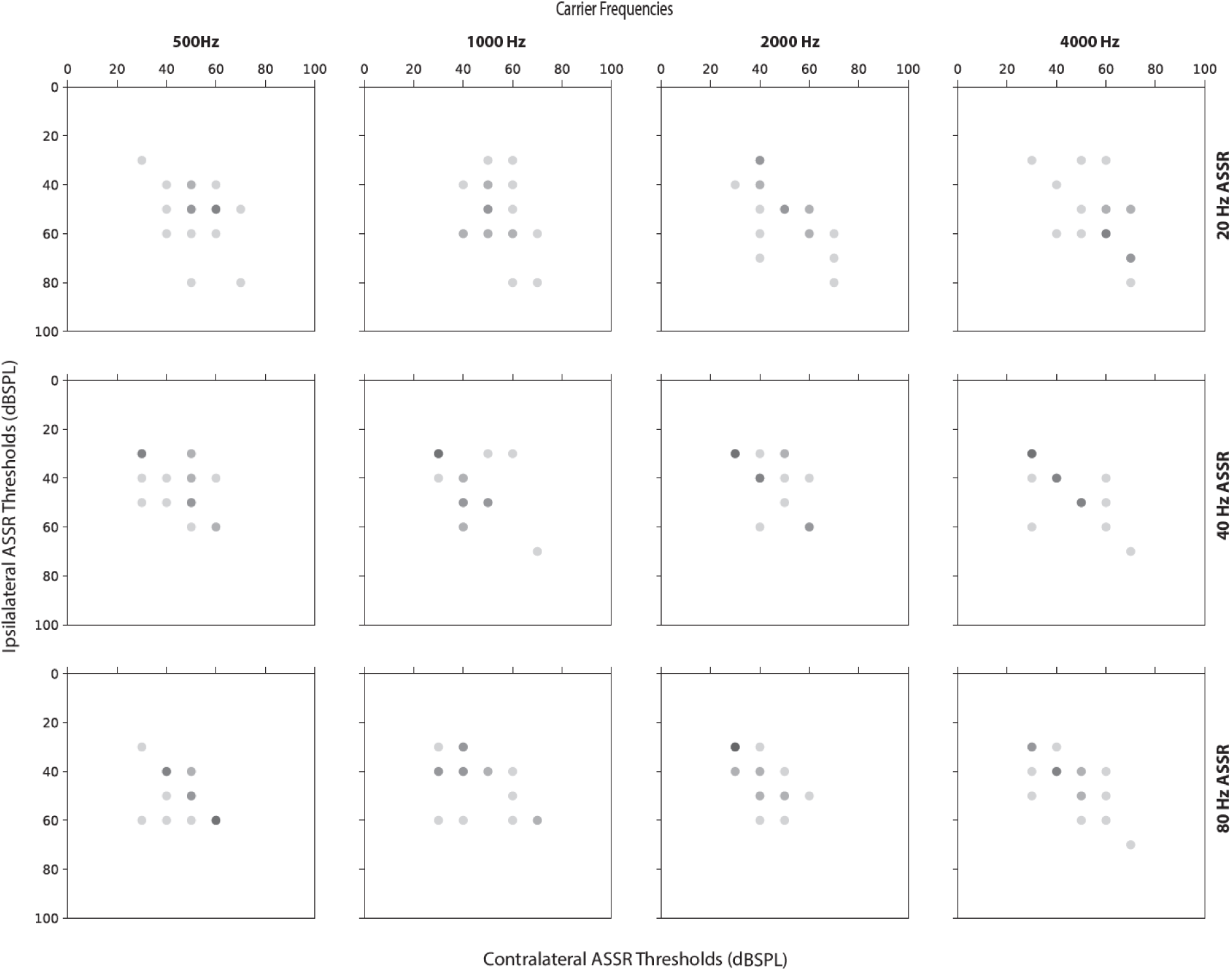
Scatterplots represent the acquired auditory steady state response (ASSR) thresholds for ipsi- and contralateral ear to the tinnitus ear. Each circle represents measurement from one subject at that combination of MF and CF. Each row represents measurements for one of the modulation frequencies 20,40, and 80 Hz. The corresponding carrier frequencies 500, 1000, 2000, and 4000 Hz are shown at the top of each panel. the darkness of the color in each circle represents overlapping datapoints (n=19 subjects).

**Table 3:** Statistical analysis results (z statistics and P-value) for comparing ipsi-vs contralateral thresholds measured with auditory steady state responses (ASSRs) in dBSPL based on the tested modulation frequency (MF) and carrier frequency (CF) using Wilcoxon signed rank test (n=19 patients).

|  |  |  | Carrier Frequency |  |  |  |
| --- | --- | --- | --- | --- | --- | --- |
|  |  |  | 500 Hz | 1000 Hz | 2000 Hz | 4000 Hz |
| Modulation Frequency | 20 Hz | Z | -0.47 | -0.25 | -0.28 | -1.33 |
|  |  | p-value | 0.64 | 0.80 | 0.8 | 0.18 |
|  | 40 Hz | Z | -0.73 | -0.57 | -1.3 | -0.18 |
|  |  | p-value | 0.46 | 0.57 | 0.2 | 0.8 |
|  | 80 Hz | Z | -1.19 | -0.53 | -1.15 | -0.51 |
|  |  | p-value | 0.24 | 0.6 | 0.25 | 0.61 |

Next, we sought to compare the obtained ASSRs in tinnitus patients and the no-tinnitus control group. Figure 2 represents box plots showing the distribution of the thresholds in each group of subjects for all MFs and CFs. The statistical analysis results are reported in Table 4. The distribution of the 20 HZ ASSR thresholds for tinnitus subjects showed slightly elevated thresholds in tinnitus patients compared to control groups (Fig. 2 top row). For instance, the estimated average ASSRs at 2000 Hz in the right ear in tinnitus patients was 47.9 ± 12.7 dB SPL compared to 43.75 ± 11 dB SPL in the control group. Similarly in the left ear at the same carrier frequency, tinnitus group showed an average ASSR threshold of 53.16 ± 13.35 dB SPL which was higher than the obtained threshold for the control group (mean ± SD: 45.4 ± 11.8 dB SPL). Such findings were consistently observed across various CFs (Fig. 2, top row). However, despite such differences, at this modulation the ASSRs between the two groups showed no significant difference (Kruskal Wallis test; p-value>0.025 in both right and left ears comparisons, see Table 4 for details).

**Fig. 2.**
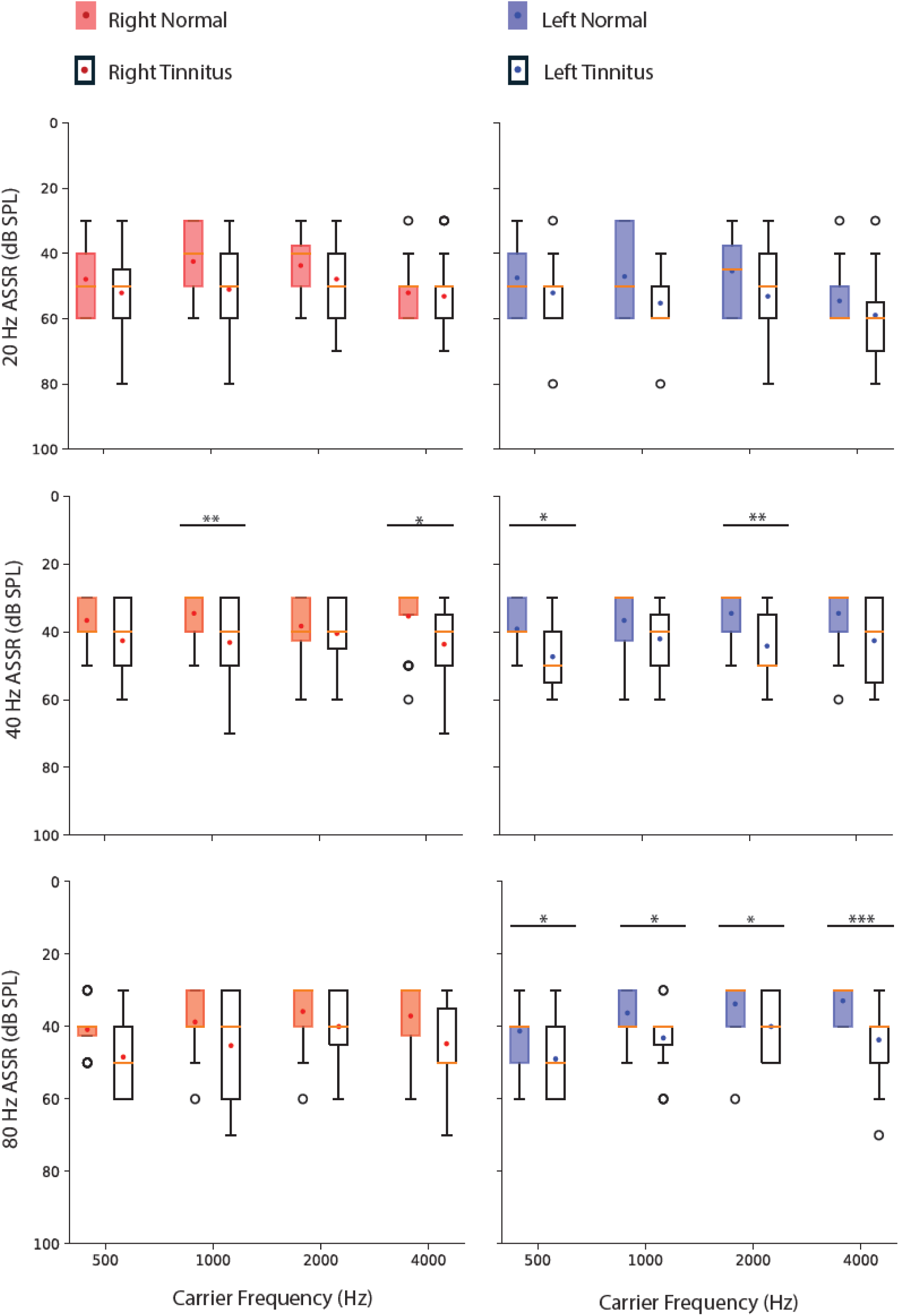
Boxplots represent the distribution of the thresholds for auditory steady state response (ASSR) thresholds for 500, 1000, 2000, and 4000 Hz. The tested modulation frequency is represented on the left side of each row. Each single boxplot represents the distribution of the thresholds between the first and third quartile, whiskers represent minimum, and maximum of the data and outliers are shown as a dot at the bottom or top of each boxplot. (n=19 subjects, for all comparisons *p<0.05, **p<0.01, ***p<0.001)

**Table 4:**
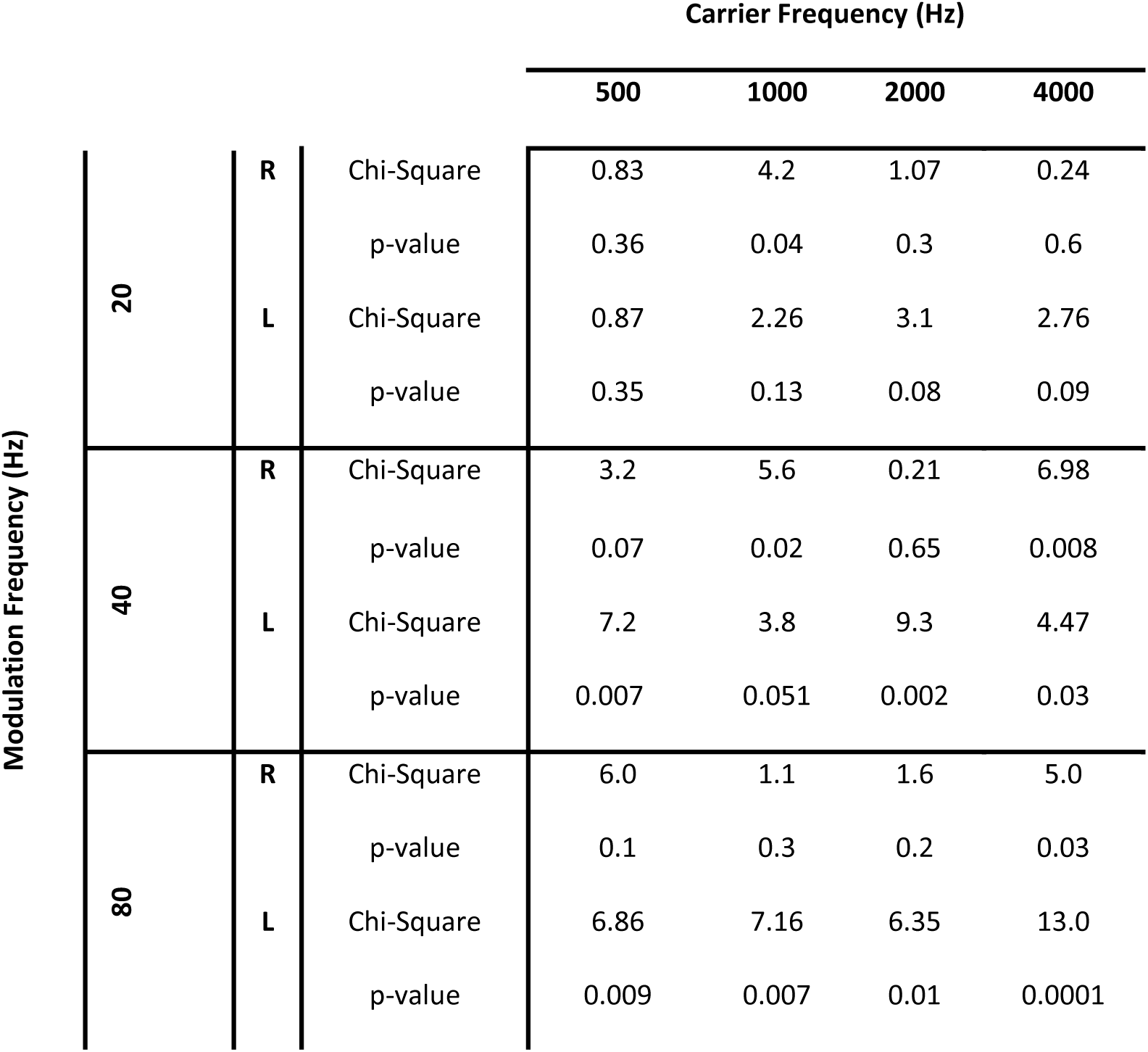
Statistical analysis results (Chi-Square and P-value) for comparing estimated right and left ASSR thresholds (dBSPL) between idiopathic tinnitus and normal hearing-no tinnitus control groups using Kruskal Wallis test (n=19 patients). Significant p-values< 0.025.

|  |  |  |  | Carrier Frequency (Hz) |  |  |  |
| --- | --- | --- | --- | --- | --- | --- | --- |
|  |  |  |  | 500 | 1000 | 2000 | 4000 |
| Modulation Frequency (Hz) | 20 | R | Chi-Square | 0.83 | 4.2 | 1.07 | 0.24 |
|  |  |  | p-value | 0.36 | 0.04 | 0.3 | 0.6 |
|  |  | L | Chi-Square | 0.87 | 2.26 | 3.1 | 2.76 |
|  |  |  | p-value | 0.35 | 0.13 | 0.08 | 0.09 |
|  | 40 | R | Chi-Square | 3.2 | 5.6 | 0.21 | 6.98 |
|  |  |  | p-value | 0.07 | 0.02 | 0.65 | 0.008 |
|  |  | L | Chi-Square | 7.2 | 3.8 | 9.3 | 4.47 |
|  |  |  | p-value | 0.007 | 0.051 | 0.002 | 0.03 |
|  | 80 | R | Chi-Square | 6.0 | 1.1 | 1.6 | 5.0 |
|  |  |  | p-value | 0.1 | 0.3 | 0.2 | 0.03 |
|  |  | L | Chi-Square | 6.86 | 7.16 | 6.35 | 13.0 |
|  |  |  | p-value | 0.009 | 0.007 | 0.01 | 0.0001 |

When comparing thresholds at 40 and 80 Hz MFs, we found consistently elevated ASSR thresholds in tinnitus subjects compared to control (Fig. 2). At 80 Hz, these differences in most carrier frequencies were more significant in the left compared to the right ear (See Table 4 for the statistical analysis results).

However, at 40 Hz ASSR we observed significantly higher ASSR thresholds in both right and left ear. Even though, the higher ASSRs were consistently seen in all CFs, this difference was only significant at some CFs in right or left ear (See Table 4 for details).

## DISCUSSION

The current study examined multiple ASSR thresholds in idiopathic tinnitus patients and normal hearing control with no tinnitus at MFs 20, 40, and 80 HZ and CFs of 500-4000 Hz. Given that ASSRs with different modulation frequencies could originate from different cortical and subcortical regions, this technique allowed us to assess the sensory processing along the auditory pathway from brainstem to cortex. Our analysis demonstrated enhanced ASSR thresholds in most CFs at 40 and 80 Hz MF corresponding to brainstem, midbrain and cortex. We further observed that the laterality of tinnitus did not affect these responses significantly. Considering both tinnitus patients and no-tinnitus control had hearing thresholds within the normal range, we believe that the enhanced responses in idiopathic tinnitus subjects could indicate possible involvement of these regions in tinnitus perception.

### The effect of tinnitus on ASSRs with different modulation frequencies

The current study aimed at assessing the changes in ASSRs with different MFs in patients with idiopathic tinnitus. Analyzing the obtained ASSRs at 80 Hz MF showed significantly enhanced thresholds in most CFs in these patients compared to no-tinnitus control. As the main contributors to ASSRs at MFs higher than 60 Hz are auditory brainstem nuclei such as cochlear nucleus and inferior colliculus [29–31], the enhanced responses at 80 Hz could point to disturbed sensory processing in these regions. Consistent with our findings, several studies have shown abnormal ABR measurements related to wave III and V in tinnitus subjects [32–35]. For instance, Said [35] compared the ABR wave latencies between sensory-neural hearing loss (SNHL) patients with and without tinnitus, and observed enhanced latency of III-V peaks for tinnitus patients compared to no tinnitus control group. Since this study used subjects with SNHL in both groups, the longer wave V latency could not be the result of hearing loss and is most likely due to the pathophysiology underlying tinnitus [35]. This was further proven in tinnitus studies with no hearing deficits [34]. Considering that the origin of wave III and V ABR are cochlear nucleus and lateral lemniscus [34, 36, 37], these results support our findings of significantly enhanced 80 Hz steady state responses in tinnitus group.

Our analysis of 40 Hz ASSRs also proved elevated responses in tinnitus patients. The ASSRs elicited at this modulation rate are proven to be mainly generated at the primary auditory cortex, midbrain, and thalamus [30,38–41]. Given the difficulty in separating these sources when recording steady state responses, it is safe to consider the involvement of either or all these regions in abnormal sensory processing in idiopathic tinnitus. Several studies using other electrophysiological or imaging approaches have previously shown the elevated midbrain or cortex responses among tinnitus patients [42]. For instance, functional changes in auditory cortex have been observed in MRI [5,44,45] and PET studies [45] show changes in the auditory cortex activity in symmetric or asymmetric tinnitus. Furthermore, changes in grey matter in thalamic regions in tinnitus subjects are also reported [46]. Other ASSR studies have shown the elevated ASSR amplitudes regardless of carrier frequencies in auditory regions based on distress level experienced from tinnitus [47]. In the current study, we only included high distress patients with a VAS score higher than 7. Thus, observing high amplitude steady state responses at the measured MFs are not surprising.

Some studies have suggested that the changes in steady state response in tinnitus patients are related to attention [48]. In this study, we deviated all participants’ attention during 20 and 40 Hz ASSR by performing simple math problems. As both tinnitus patients and control group were similarly engaged in a task at lower MFs, we believe the changes in ASSR thresholds were not due to attention. This is further supported by another study which used N1 wave in magnetoencephalography (MEG) to investigate the effect of attention in tinnitus patients and found no attention-induced effect on N1 wave in these patients [49].

We observed different effects on various CFs in 40 and 80 Hz ASSR with some CFs being more affected than the others in tinnitus subjects. As we were not able to identify the tinnitus frequency in the patients used in this study, it is not clear whether the differences between tinnitus patients and the control group at certain CFs were due to the interference with tinnitus sensation at a particular frequency. Studies on tonotopic organizations of 40 Hz ASSR have shown that with increase in the CF, the neural generators of ASSRs shift laterally on the auditory cortical regions [41]. In the current study we observed changes in ASSR thresholds in tinnitus patients compared to control to be affecting some CFs but not others. Considering that not all affected CFs were adjacent frequencies (1000 and 4000 Hz in right vs 500 and 2000 in left) this might suggest a distribution in some tonotopic regions that could gradually spread to other tonotopic regions.

Our findings did not show any major difference in the obtained 20 Hz ASSR thresholds for idiopathic tinnitus and no-tinnitus control. Overall, the obtained thresholds in this MF were more enhanced compared to other MFs in both groups of tinnitus patients and control. This could be the result of challenges in creating a reliable EEG signal above the background noise at this MF [50] reported by different studies, which creates challenges in localizing the origin of these responses with high certainty [20].

### Steady state responses and tinnitus laterality

Previous research has shown contradictory findings in comparing neuronal responses based on tinnitus laterality. For instance, fMRI studies on tinnitus subjects have previously shown abnormal IC activation in the side contralateral to the tinnitus percept side [11], while others have seen no asymmetry in activation pattern of this region in lateralized tinnitus patients [51]. Our findings support the latter, as we did not observe any significant differences between the thresholds acquired in any combination of MF and CF between the ipsi- and contralateral ASSR thresholds. The differences in the findings here and in previous studies could be further related to the tinnitus population under study. In the current study, we applied strict exclusion criteria to limit the population under study to those with idiopathic tinnitus. This removes many underlying factors such as SNHL that could contribute to tinnitus perception and laterality.

### Conclusion

Tinnitus is suggested to be the result of an imbalance between inhibitory and excitatory processes in the midbrain, brainstem, and auditory cortex. These are the auditory brain regions where neural activity from somatosensory and auditory system interact. It has been proposed that such imbalance causes hyperexcitability that results in tinnitus perception [52]. Various medications have been used for alleviating tinnitus perception by targeting the underlying cause. Since effectiveness of such interventions largely relies on understanding the underlying cause of tinnitus, localizing auditory regions with sensory processing deficits in idiopathic tinnitus is crucial. Findings of this study supports involvement of multiple regions along the auditory pathway in these patients which could be the results of malfunction of a distributed network involving various brain areas in creating tinnitus sensation [20].

## Data Availability

All data produced in the present study are available upon reasonable request to the authors.

## Note

### Conflict of Interest

The authors declare that they have no conflict of interest.

## Notes

### Competing Interest Statement

The authors have declared no competing interest.

### Clinical Protocols

https://pubmed.ncbi.nlm.nih.gov/31115686/

### Funding Statement

This study did not receive any funding.

### Author Declarations

Ethics committee/IRB of Iran University of Medical Sciences and Tehran University of Medical Sciences gave ethical approval for this work.

